# Grand battement kinematics and aesthetics in adolescent recreational dancers: Short term effects of static and dynamic hamstrings stretching

**DOI:** 10.1101/2020.09.15.20195065

**Authors:** Frédéric Dierick, Fabien Buisseret, Loreda Filiputti, Nathalie Roussel

## Abstract

We have studied the kinematics of ballet figures performed by adolescent recreational dancers and determined the most effective muscle stretching modality allowing to increase their physical performance while not harming the aesthetic perception of their motion. Sixteen participants aged between 10 and 19 years were recruited to perform a grand battement, before and after a static or dynamic stretching of hamstring muscles. The three-dimensional kinematics of the grand battement was measured by an optoelectronic system and the aesthetics was scored from a video watched by a jury of professional dancers. Our results show that stretching has a significant impact on grand battement kinematics, and that the most important modifications are induced by dynamic stretching rather than static stretching. Dynamic stretching significantly improves the explosive aspects of the movement (duration and maximal speed), but leads to a significant reduction of its smoothness (jerk). Significant correlations between kinematic parameters and aesthetic scores have been observed, one of them being a positive correlation between thigh’s range of motion and total aesthetic score. These correlations can serve as a reference for movement analysis experts, dancers and their teachers to improve physical performance required by the standards of today’s dance practice without altering the corresponding aesthetic judgment.

## I. INTRODUCTION

Dance is a form of self-expression in which dancer’s artistic intentions and the laws of biomechanics are intermingled. Dancers physical skills condition their ability to reach their aesthetics ambitions. Hence, during the execution of figures, dancers have to adopt an optimization of their kinematics on the basis of a trade-off between physical skills (such as range of motion (ROM), speed of execution) and aesthetic self-expression (movement’s beauty/likeability/pleasantness), the latter relying on personal or external criteria (judgment of jury or audience).

One of the first dancing masters who defined the bases of the classical technique was Pierre Beauchamps (1631-1705). Since the practice of dance is associated with firmly established traditions, transmitted mainly through teaching. However, these traditions are not entirely immutable. For example, over the last century, it has been noted that dancers were required to demonstrate increasingly greater ROMs, with aesthetic judgment favouring the latter [1]. As a result, today’s dancers are subject to more physiological constraints than their predecessors to practice their art.

It is nowadays relevant to study the kinematics of the figures performed by adolescent recreational dancers in detail since: (1) faulty kinematics during the execution of dance movements may be associated with the development of neuro-musculoskeletal injury; (2) high dance-related neuro-musculoskeletal injury incidence rates were reported in adolescents. For example, dancers with Achilles tendinopathy demonstrate increased peak transverse and frontal plane kinematics when performing the takeoff of a saut de chat compared to a control group [2]. Also, reported injuries assessed by a physiotherapist in adolescent dancers at a liberal arts high school dance program range from 2.6 (females) to 5.5 (males) injuries per 1,000 dance hours and the most common locations for injuries were ankles, lower leg/calf, and back, usually caused by overuse, muscle strains, and sprains [3].

As noticed by observation of habits in dance schools, static muscle stretching is nowadays mostly used by recreational dancers to increase the ROM allowed by the joints. However, there are various muscle stretching methods: static, dynamic, and ballistic [4, 5]. According to these studies, dynamic muscle stretching seems to be relevant in the of reactive activities, i.e. activities that require explosiveness, strength and speed. Such activities exist in classical dance movements such as in high velocity kicks (grand battement). In this study, we consider the impact of two muscle stretching methods, static and dynamic, on the aesthetic and kinematic performance of grand battement in adolescent recreational dancers.

It is particularly important to determine the best stretching practices in recreational adolescent dancers. They are in a period of their life characterized by significant biological and psychological changes. They often train many hours, but they still do not have a fully mature skeleton. As skeletally immature individuals are more vulnerable to trauma due to explosive muscle contractions [6, 7], finding the most the most effective muscle stretching modality is of great importance. It should be able to increase the physical skills while not harming, or even improving, the aesthetic self-expression performance. The aim of this study is therefore twofold. First, to evaluate the impact of different stretching methods on the kinematics of the grand battement. Second, to highlight the existence of correlations between lower limb kinematic parameters and the perception of the aesthetics of the movement assessed by a jury. Although the variables involved are not a priori known, it is tempting to state that kinematics and aesthetics must be correlated as shown in professional dancers [8, 9].

## II. MATERIAL AND METHODS

### A. Dancers and jury members

Sixteen participants were recruited from two private dance schools around Charleroi (Belgium) and from our Physical Therapy Department at Haute Ecole Louvain en Hainaut. A written informed consent was signed by each participant or by his/her legal guardian if he/she was younger than 18 years after being informed of the experimental protocol of the study and the use of their personal data. The experimental protocol has been accepted by the local internal commission and respects the Helsinki Declaration on Ethical Principles for Medical Research Involving Human Beings. The protocol of the experiment has been approved by the Academic Bioethics Committee (n° B200-2019-161).

To be included in the study, participants had to be recreational adolescent dancers, *i*.*e*. aged between 10 and 19 years [10]. The latter age range is the most represented in dance schools. Participants had to know the basics of classical/contemporary dance and had to be trained at performing standard figures such as grand battement and développé. They had to train at least 3 hours per week. Participants with a history of lower limb or spinal trauma during the 6 months preceding the data acquisition, whether or not it required physiotherapy, medical or surgical management were excluded.

A jury of five professional dancers was chosen among personal acquaintances in order to judge the aesthetics of the figures performed by the participants. They had all had classical dance training and had been giving dance lessons for at least one year. The members of the jury were blinded to the medical and artistic background of the participants and signed a confidentiality agreement concerning the performances they were asked to watch.

### B. Experimental protocol and data acquisition

Each participant was asked to perform a grand battement from first position in three conditions: control (CRTL), after static stretching (SS) and after dynamic stretching (DS). CTRL condition always came first, and the other two conditions were randomly chosen. The same experienced operator (LF) was in charge of the protocol realization.

#### CTRL condition

Participants performed a grand battement from first position without prior hamstring muscles stretching. They received the following instruction: “Make your best movement, as you were taught.”

#### SS condition

A static hamstring muscles stretching posture was performed for 30 seconds, which is the time needed to efficiently stretch the muscles [11]. It was performed twice on both lower limbs with maximum passive hip flexion and knee extension ROM below the pain threshold. For this purpose, participants were instructed to experience the maximum stretching sensation without any burning, tearing, tingling, or shaking sensation. For the stretching posture execution, the foot heel was placed on a wall so that the maximal amplitude of hip and knee joints was reached (Fig. 1). Then the participant was instructed to bend the trunk to increase the stretch. Another proposal for less flexible participants was to place the foot heel on a table and perform a trunk bending. Another grand battement was performed immediately after static stretching postures with the same instruction as in CTRL condition.

**FIG. 1:**
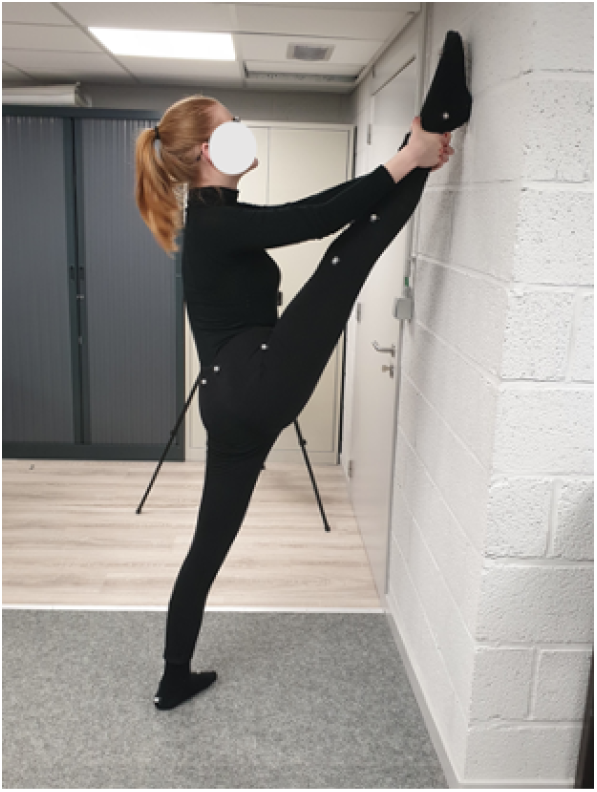
The static hamstring muscles stretching posture: Picture of one participant in the laboratory where measurements were performed.

#### DS condition

A dynamic hamstring muscles stretching movement was performed at high velocity on both lower limbs. It consisted of throwing the lower limb forward with the knee extended. It was instructed to the participants to restrict the abduction movement of the hip to accentuate the stretching of the hamstring muscles. Ten beats of each lower limb were performed at a frequency of approximately 1Hz so that all subjects followed the same rhythm marked by a metronome. Each movement had to cover the entire active ROM of the participant’s hip joint [5]. Another grand battement was performed immediately after dynamic stretching movements with the same instruction as in CTRL condition.

A 15-minute break was imposed between two consecutive conditions, with CTRL condition first and SS and DS conditions randomized after. The 15-minute break is more than the 10 minutes found to be necessary in [12] for the muscle to return to its initial state.

Recording of the kinematics of the grand battement was performed once per condition, unless participant declared the motion unsatisfactory. Prior to CTRL condition, participants were equipped with sixteen passive reflective markers with a size of 14 *mm* in diameter allowing kinematic data to be recorded by a motion capture system (Vicon Motion Systems Ltd, Oxford Metrics, Oxford, United Kingdom) consisting of 8 optoelectronic cameras (Vero v.2.2) with a sampling frequency of 120 *Hz*. The placement of these markers, using double-sided adhesive tape, was applied according to the lower body Plug-in-Gait model (Oxford Metrics, Oxford, United Kingdom). The model includes two marker positions; twelve markers symmetrically placed on anatomical landmarks identifiable by palpation, i.e., on the anterior and posterior superior iliac spines (ASI and PSI), on the knee flexion/extension axis (KNE), on the lateral malleolus along the imaginary line passing through the trans-malleolar axis (ANK), on the head of the 2^nd^ metatarsal (TOE) and on the calcaneus (HEE). The asymmetry of the other four markers, namely those positioned at the lower third of the lateral side of the thigh (THI) and leg (TIB), was necessary for easily differentiate between the left and right side. Participants were asked to wear only shorts for men and a bra for women in order to maximize the visualization of the markers and their appropriate positioning throughout the measurements.

After the recording of the grand battements in each condition, Vicon Nexus software (v.2.7.1, Oxford Metrics, Oxford, UK) was used for reconstruction and three-dimensional (3D) modeling of markers. A Plug-in Gait Dynamic pipeline was applied with a Woltring quintic spline algorithm with a mean square error value of 10. The data were then exported with the ASCII standard in a csv file for further processing. This file included all the 3D positions of the markers, 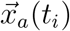 giving the three-dimensional kinematics of the marked anatomical landmarks *a* at time *t*_*i*_, as well as the value of the thigh (*θ*_*THI*_), knee (*θ*_*KNE*_), and ankle (*θ*_*ANK*_) angles as function of time. Successive time values are separated by *t*_*i*+1_ *− t*_*i*_ =Δ*t* = 1*/f*. *t*_*i*+1_ *− t*_*i*_ = Δ*t* = 1*/f*. A schematic view of the most relevant markers is shown in Figure 2 A, and a typical plot of grand battement kinematics is shown in Figure 2 B. Typical traces of angular time series during a grand battement are shown in Figure 2 C.

**FIG. 2:**
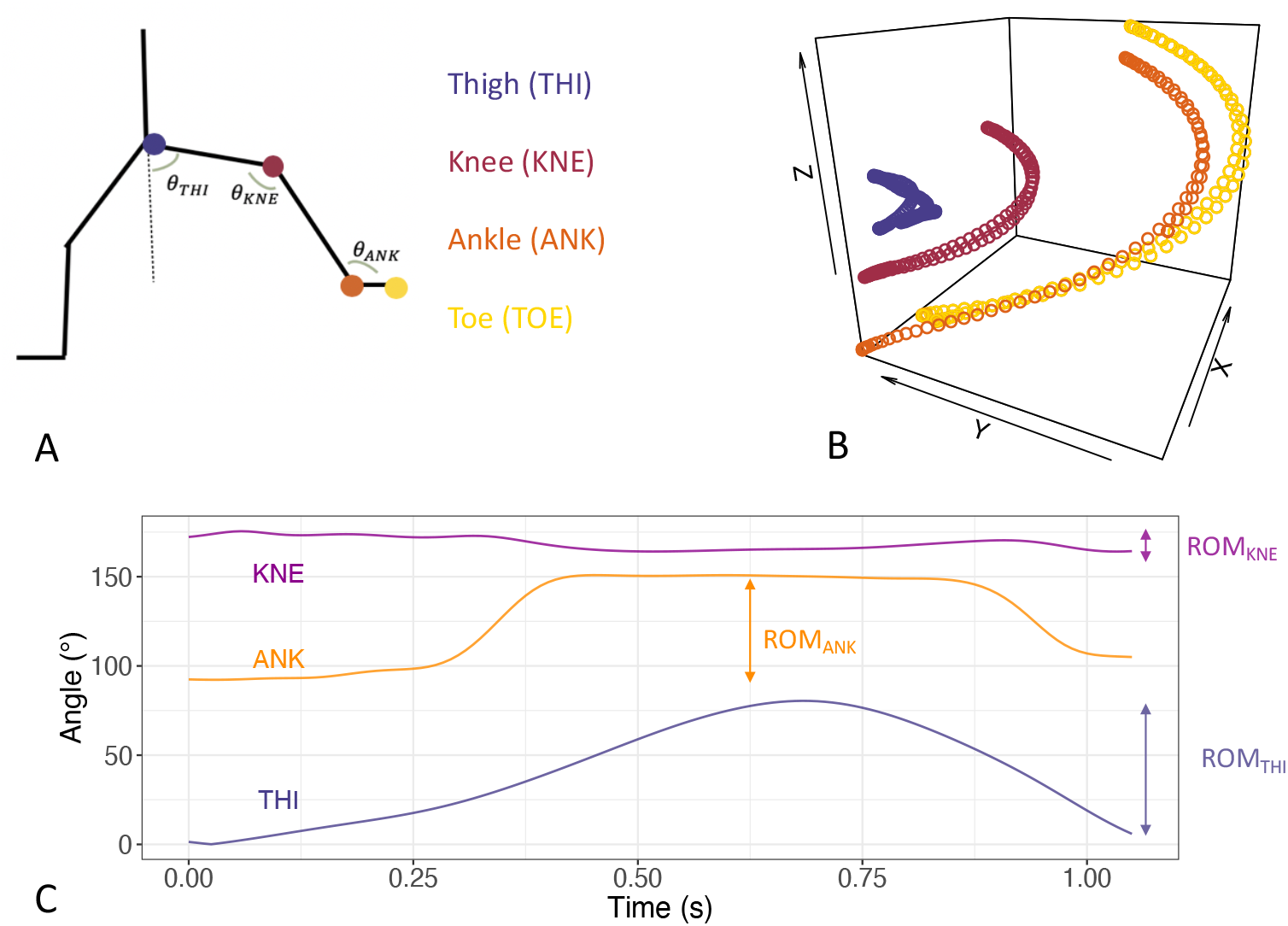
A. Four anatomical landmarks of particular interest for the study of grand battement: lower third of the lateral side of the thigh (THI), knee flexion/extension axis (KNE), lateral malleolus along the imaginary line passing through the trans-malleolar axis (ANK), head of the 2^*nd*^ metatarsal (TOE). The three angles are the angle between the thigh and the vertical (*θ*_*THI*_), the angle of the knee (*θ*_*KNE*_) and the angle of the ankle (*θ*_*ANK*_). B. Typical three-dimensional trajectory of one participant performing a grand battement. The following time series are shown: 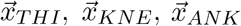 and 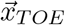 C. Typical angular trajectories during a grand battement (same participant as in B). The range of motion (ROM) is illustrated for the three angles.

In addition, the video recordings of the grand battements made by a dedicated camera (Vicon Vue) were extracted and merged in random order to be shown to the jury during a single evaluation session. The jury’s evaluation is referred to as aesthetic assessment. Each member had to rate all the 48 grand battements (16 participants × 3 conditions) after a single view of each, without talking or communicating with the other members. The members of the jury were not informed of the experimental protocol and of the different conditions. The aesthetical assessment was made by using the questionnaire presented in Table I. It was inspired from [13] but adapted to our protocol: Items related to longer-duration performances have been dropped. The questionnaire was explained to the members of the jury before the session.

**TABLE I:** Questionnaire used for aesthetic assessment of the grand battements by the jury members. The first three items are adapted from criteria 1, 3 and 4 of [13]. We have added the item4. The meaning of the score was: 1-not mastered, 2-poorly mastered, 3- satisfactorily mastered, 4-Fully mastered.

| Criterion | Description | Score |
| --- | --- | --- |
| 1. Control of movements (/8) | Controlled landing from jump/turn, controlled lifting | 1 – 2 – 3 – 4 |
|  | Core strength, alignment, posture | 1 – 2 – 3 – 4 |
| 2. Accuracy of movements (/16) | Arm and pelvis correct placement | 1 – 2 – 3 – 4 |
|  | Feet correctly placed on the ground | 1 – 2 – 3 – 4 |
|  | Fully stretched leg extensions (if required) | 1 – 2 – 3 – 4 |
|  | Pointed/stretched foot while lifting | 1 – 2 – 3 – 4 |
| 3. Technique (/4) | Elevation techniques, height of extensions | 1 – 2 – 3 – 4 |
| 4. Smoothness (/4) | Smooth trajectory of lower limb | 1 – 2 – 3 – 4 |

### C. Data analysis

The duration *T* was computed from 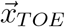 as the time necessary for the foot to leave the ground and come back. The instantaneous speeds of the different anatomical landmarks, 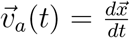, were computed from 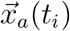by the standard discretized time derivative 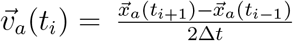 The corresponding norms *v*_*a*_(*t*_*i*_) were then computed as well as the maximal speeds

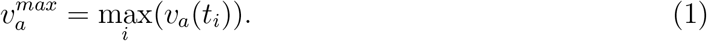

We focused on the thigh and ankle maximal speeds, denoted as 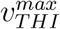 and 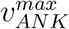 respectively. These speeds give the typical magnitude of lower limb speed and should be similar during a grand battement.

Angular ROM were computed from angular time series as *ROM*_*a*_ = max_*i*_(*θ*_*a*_(*t*_*i*_)) *−* min_*i*_(*θ*_*a*_(*t*_*i*_)), see Figure 2 C for a typical trace of the angular time series and a graphical illustration of the ROM.

The angular instantaneous speeds, 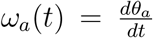, accelerations 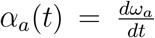, and jerks, 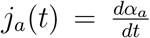 were also computed by using the same discretized derivative as for 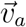 The maximal angular speeds were obtained by using 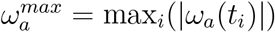

The log-dimensionless jerks *J*_*a*_ were finally computed thanks to the definition of [14]

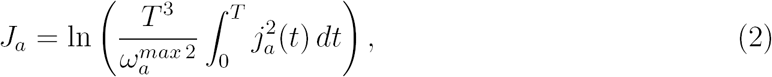

where the integral is approximated by the discretized expression ∫ *f* (*t*)*dt* =Σ_*i*_ *f* (*t*_*i*_)*dt*. Log-dimensionless jerk is a valid and sensitive tool to assess motion smoothness from kinematical data, see [14] for a detailed discussion and comparison to other candidates. It is worth recalling that a high smoothness is related to a low log-dimensionless jerk.

For the aesthetic questionnaire, for each participant in each condition we gathered the total score (/32) and the technique (/4) and smoothness (/4) subscores given by each member of the jury. By summing all member’s scores we obtained a total score (/160) for a participant in a given condition as well as a technique (/20) and smoothness (/20) subscore.

### D. Statistical analysis

The influence of condition (CTRL, SS, DS) on kinematic parameters was assessed by a one-way analysis of variance with repeated measures (ANOVA RM). In case of significant influence of condition, the conditions were compared pairwise by a Holm-Sidak post-hoc. The influence of condition on jury’s scores was assessed by a one-way non-parametric ANOVA RM (Friedman test). Significance level was set to *p* = 0.05 and tests were performed by using Sigmaplot software (v.11.0, Systat Software, San Jose, CA).

The agreement between members of the jury was assessed by computing Kendall’s *τ* correlation coefficient for all pairs of members. Correlations between kinematic parameters and aesthetics scores were studied by computing Pearson’s correlation coefficients (*r*), regardless of condition. We consider a correlation as significant if |*τ* | or |*r*| *>* 0.3 and *p ≤* 0.05 following Guilford lines [15]. Further analysis of correlations between variables was made by performing a partial component analysis (PCA) on all computed variables, regardless of condition. PCA and correlation coefficients computations were performed with R software (v.3.4.2), packages *corrplot* and *FactoMineR*.

## III. RESULTS

### A. Dancers and jury members

A description of the main characteristics of the dancers and jury members is available in Table II.

**TABLE II:** Dancer’s (left) and jury member’s (right) main characteristics. BMI: body mass index.

|  | Dancers | Jury |
| --- | --- | --- |
| N | 16 | 5 |
| ♂/♀ | 4/12 | 1/4 |
| Age (years) | 15.3±2.8 | 25.8±4.1 |
| Height (cm) | 161±12 | 164±4 |
| Body mass (kg) | 56.4±14.6 | 57.8±5.8 |
| BMI (kg/m<sup>2</sup>) | 21.4±3.6 | 21.3±1.1 |
| Dancing from (years) | 7.5±3.8 | 18.6±4.6 |
| Training (h/week) | 8.8±4.3 | 14.8±8.5 |

### B. Kinematics and aesthetics

Mean*±* standard deviations values of the computed kinematic parameters during grand battement are given in Table III for the different conditions. Several parameters are significantly modified by the condition: 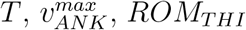, and *J*_*THI*_. As shown by the post-hoc analysis results in Table IV, the observed differences are mainly due to the differences between CTRL and DS condition. When the SS condition is significantly different than the CTRL condition, it cannot by distinguished from the DS condition. After DS, the grand battement is executed faster (smaller *T* and larger 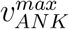 values) and with a larger ROM (larger *ROM*_*THI*_ value) than in the CTRL condition. The grand battement is moreover less smooth after DS (larger *J*_*THI*_ values). These results are displayed in Figure 3.

**TABLE III:**
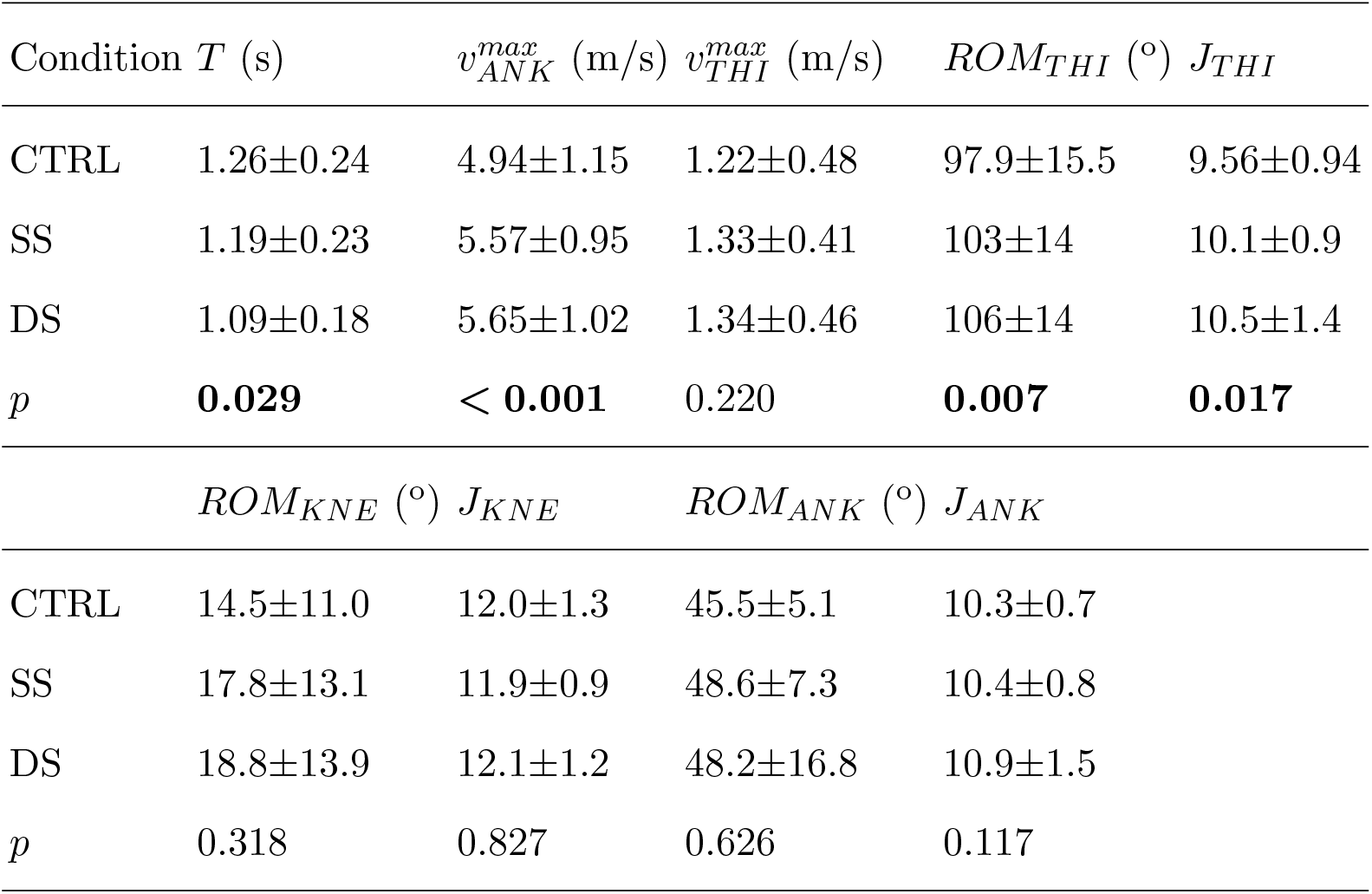
Results of the one-way ANOVA RM performed on the computed kinematic parameters during grand battements execution in the different conditions. Significant differences between conditions are written in bold font. Results are given under the form mean*±* standard deviation.

**TABLE IV:** Post-hoc analysis results. *p−* values are given for condition comparisons. Significant differences between conditions are written in bold font.

| Comparison | $T$ | $v_{ANK}^{max}$ | $ROM_{THI}$ | $J_{THI}$ |
| --- | --- | --- | --- | --- |
| CTRL vs SS | 0.247 | < **0.001** | 0.069 | 0.219 |
| CTRL vs DS | **0.026** | < **0.001** | **0.007** | **0.014** |
| DS vs SS | 0.213 | 0.626 | 0.264 | 0.158 |

**FIG. 3:**
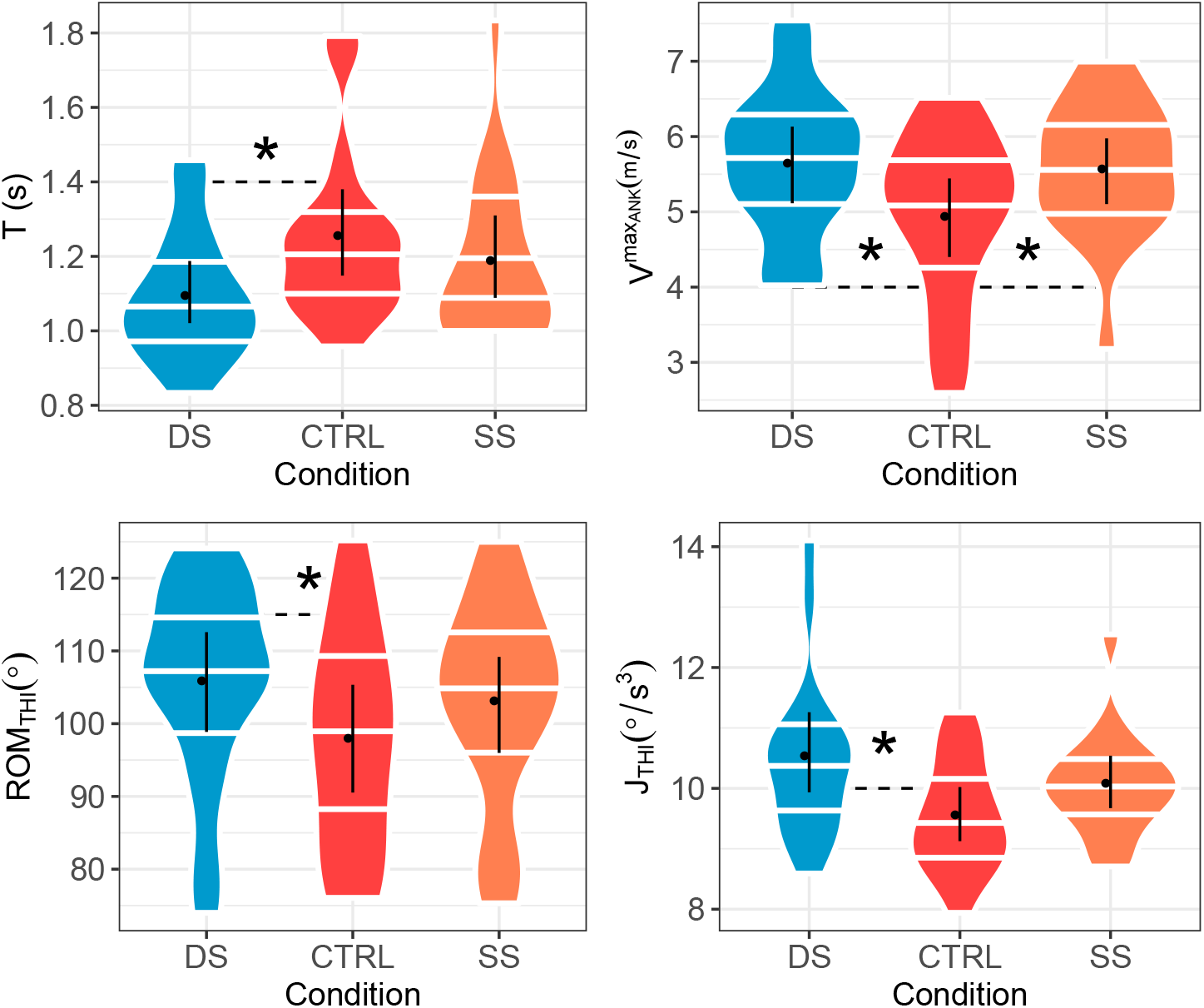
Graphical illustration of one-way ANOVA RM results for the kinematic parameters showing significant differences between conditions. The distributions of the results in the different conditions are displayed by a violin plot, with the median, first and third quartiles shown as horizontal white lines. Means and standard deviations are also shown (black points with vertical bars). Significant differences between conditions are marked by a ^*^.

A necessary condition for the aesthetic assessment to be relevant is that the different members of the jury agree on the compared performances of the different dancers. This agreement is assessed by Kendall’s *τ* coefficient, whose minimal and maximal values are given in Table V. The definition

**TABLE V:** Minimal (Min) and maximal (Max) Kendall’s *τ* coefficients between members of the jury for the technique and smoothness subscores, and for the total score. *p−*value is given between brackets. Significant correlations are written in bold font.

|  | Technique | Smoothness | Total |
| --- | --- | --- | --- |
| Min | **0.31(0.01)** | 0.24 (0.03) | **0.37(0.01)** |
| Max | **0.59(< 0.01)** | **0.58(< 0.01)** | **0.59(< 0.01)** |

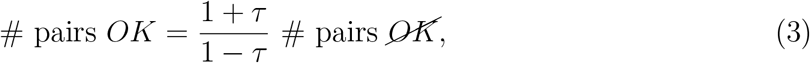

where the symbol # denotes the number of objects, allows the following interpretation of the results: Members of the jury agree on the hierarchy of a given pair of performances about 2 to 4 times more than they disagree. The best agreement is reached for the total score and for the technique subscore. As shown in Table VI, aesthetic assessment is not significantly dependent of the condition.

**TABLE VI:** Results of the Friedman test performed on the jury’s technique and smoothness sub-scores, and on the total score. Results are given under the form median [*Q*1 *− Q*3].

| Condition | Technique (/20) | Smoothness (/20) | Total (/120) |
| --- | --- | --- | --- |
| CTRL | 14 [13 – 15] | 14 [12 – 15] | 120 [104 – 137] |
| SS | 15 [13 – 17] | 15 [12 – 16] | 123 [117 – 137] |
| DS | 14 [13 – 17] | 14 [12 – 15] | 128 [97 – 139] |
| $p$ | 0.630 | 0.820 | 0.872 |

### C. Aesthetics-kinematics correlations

Pearson’s correlation coefficients between the aesthetic assessment and the kinematic parameters are given in Table VII. The largest positive correlations are observed between the scores and 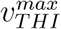 and *ROM*_*THI*_: grand battements with high thigh speed and large thigh ROM have better scores. On the contrary negative correlations are observed between the scores and *ROM*_*KNE*_ and *J*_*ANK*_. Two examples of correlated measurements are shown in Figure 4.

**TABLE VII:**
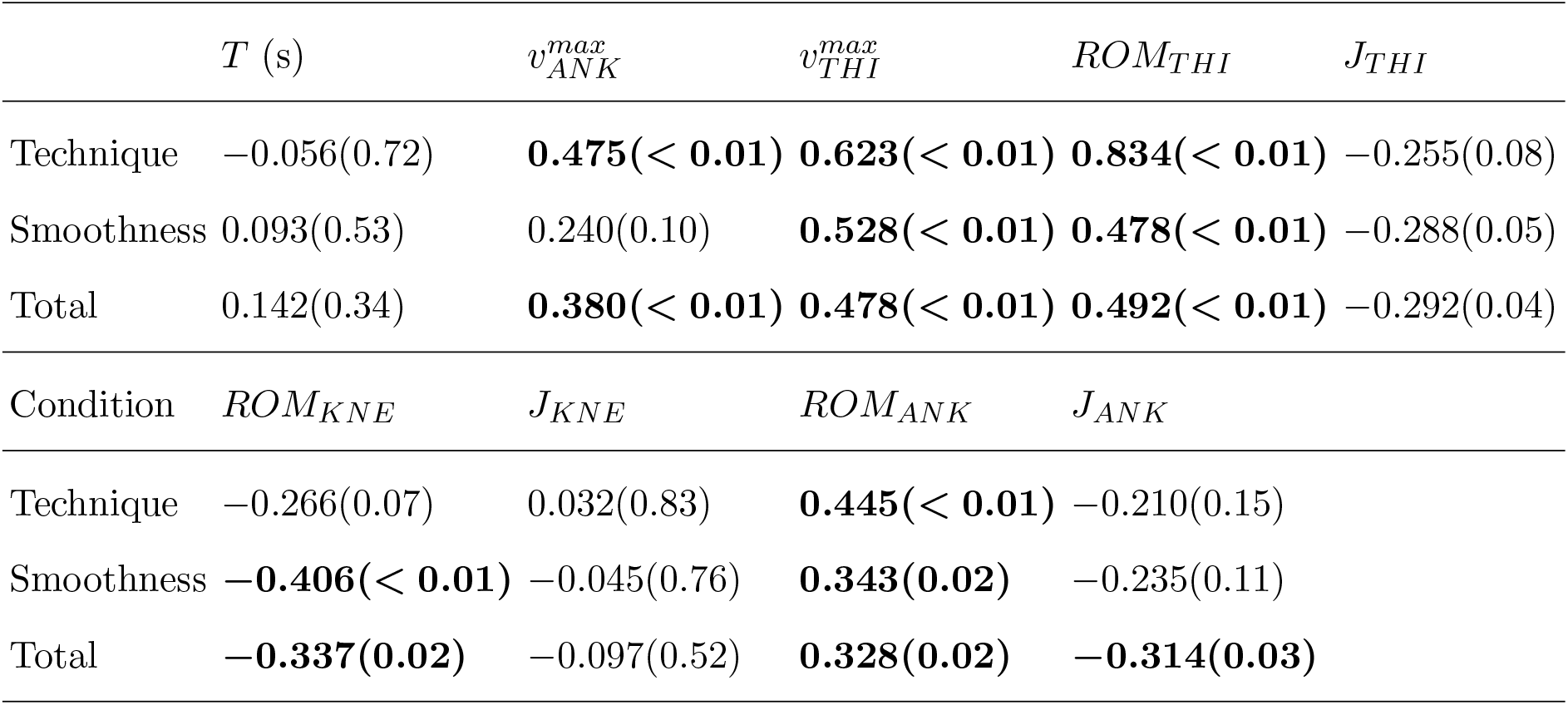
Pearson’s correlation coefficient between jury’s scores (technique, smoothness and total) and kinematic parameters. *p−*values are given between brackets. Significant correlations are written in bold font.

**FIG. 4:**
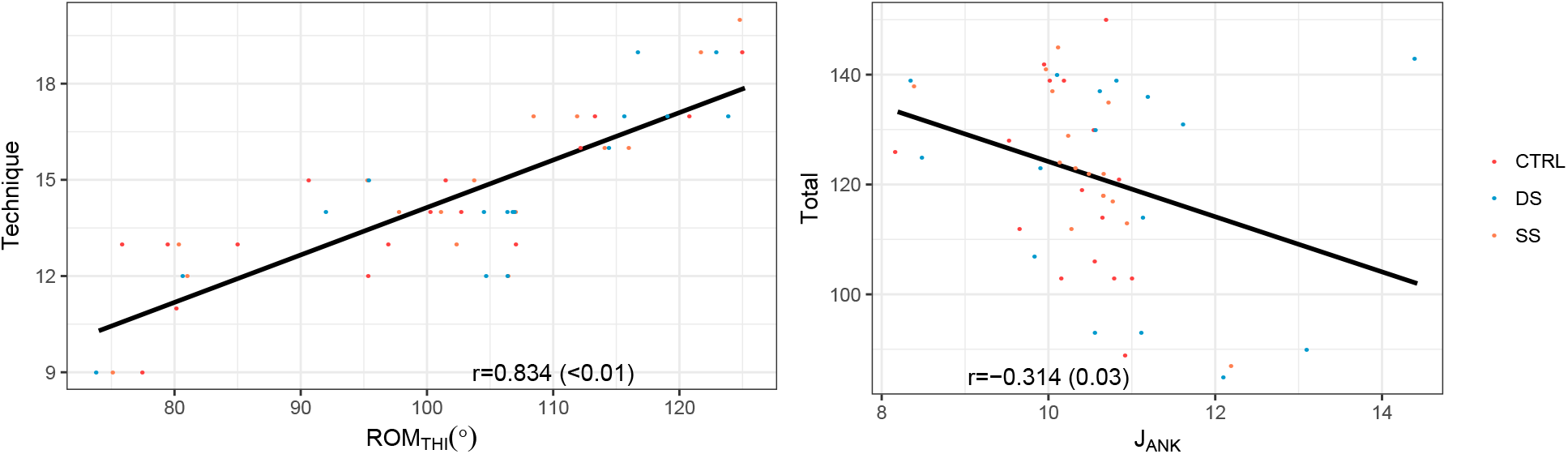
Two examples of significant correlations between jury’s scores and kinematic parameters. Left: The best observed correlation, between the technique subscore and *ROM*_*THI*_. Right: The poorest observed correlation, between the total score and *J*_*ANK*_.

The PCA displayed in Figure 5 shows that the set of computed parameters accounts for 69.4% of the total participant’s variability including the first three dimensions. The correlations between the scores and thigh and ankle maximal velocity and ROM appear clearly along the first dimension. The kinematic parameters with the maximal contribution along the first dimension are 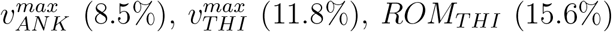. Jerk-related quantities have the maximal contribution to the second dimension: *J*_*ANK*_ (8.7%), *J*_*THI*_ (24.7%), *J*_*KNE*_ (29.8%). Grand battement’s duration is dominant in the third dimension (36.6%).

**FIG. 5:**
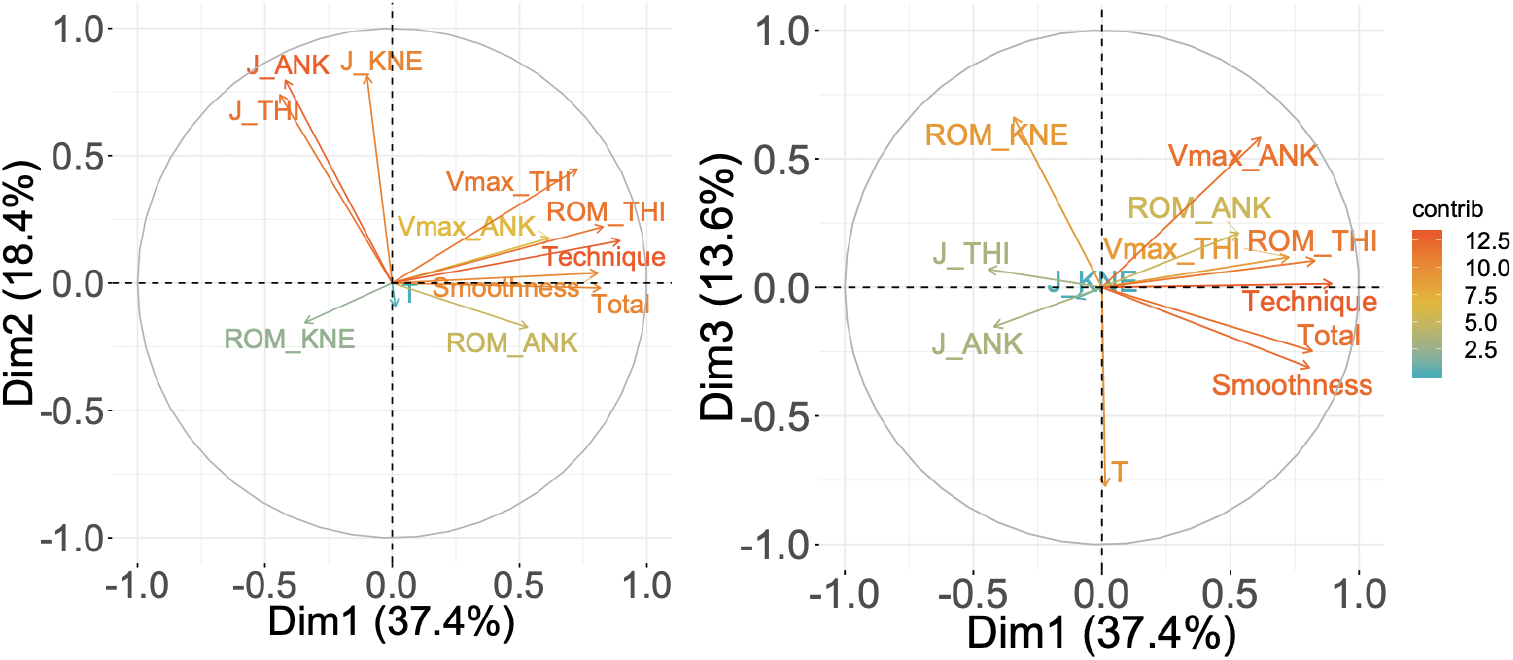
PCA performed on the computed parameters. The correlation circles for dimensions 1 and 2 (left panel) and dimensions 1 and 3 (right panel) are shown. The contribution of each variable to the principal axes (contrib) are coded in colors, with cold colors (turquoise blue) showing low contribution and warm colors (orange) high contribution.

## IV. DISCUSSION

To our knowledge, this is the first study assessing the short term effects of SS and DS of hamstring muscles on grand battement aesthetics and kinematics in adolescent recreational dancers. Our results show that stretching has a significant impact on grand battement kinematics, and that the most important modifications are induced by DS.

The positive impact of DS on ROM around a joint could be attributed to reduced stiffness of the muscle-tendon unit and to the improved muscular performance subsequent to temperature increase and postactivation potentiation mechanism [16], a transient improvement of muscular contractility following a voluntary contraction [17]. Unfortunately, we have not made electromyographic measurements so we cannot confirm stretching-induced modifications on muscular activation patterns as in [18] on jump performance. The amplitude of grand battement at thigh (see ROM_*THI*_) is significantly larger after DS in coherence with the findins of [19]. It is an important result because larger amplitudes are correlated with a better aesthetic score. Grand battement is also faster after DS: the duration of execution (*T*) is smaller and the ankle maximal speed 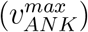 is larger. It has already been hypothesized that DS induces larger muscular forces, which in turn caused larger accelerations and maximal speeds [20]. SS has a positive impact on maximal speed too but its impact on grand battement execution was more limited.

The first two dimensions of the PCA allow us to separate the kinematic parameters into two separate categories: “performance” or physical skills (ROM and maximal speed) and “smoothness” (jerk). Performance parameters are positively correlated with the total score and with the technique and smoothness subscores. The correlation between maximal speed at hip 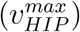 and ankle 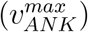 and total score is an expected consequence of ballet rules: The grand battement has to be an explosive ballet figure, therefore the jury favors the fastest executed figures. The positive correlation between ROM_*THI*_ and total score is coherent with the results obtained in the study of [1]: large ROM in dance are favored by the audience. It has to be noted that jerks (*J*_*ANK*_, *J*_*KNE*_, and *J*_*THI*_) are not significantly correlated with smoothness subscores as we would have thought a priori. The concept of kinematic smoothness is probably more related to complex movements and longer performances in jury’s perception; it is likely that professional dancers are not “trained” to assess kinematic smoothness as accurately as they can perceive speed and ROM. Kendall’s *τ* coefficients confirm the latter assumption: The agreement between members of the jury is better for the total score and the technique subscore than for the smoothness subscore.

Correlation between biomechanical and aesthetic perceptual variables have already been observed in dance figures [8]. However, perceptual ratings of dance are specific to both the task and the context [9]. The task is here the grand battement, and the context is that of a laboratory. It can be thought as a limitation of the present study: To what extent our conclusions would apply to on stage performances remains an open question. An other limitation is that we only performed a kinematic analysis of the lower limb performing the figure. As shown in [21], pelvic motion is not negligible in battement, and although the active lower limb is dominant, a jury may also be influenced by such movements.

Although DS improves the explosive aspects of grand battement (time and speed), it has to be noted that this stretching method leads to a significantly larger *J*_*THI*_, hence to less smooth thigh movement. Our participants being recreational dancers, we think that the gain in ROM_*THI*_ they experienced because of stretching led them to a poorer control at high thigh angle and eventually to a more fluctuating velocity profile, resulting in a larger jerk [22].

Both stretching methods tend to increase *ROM*_*ANK*_, but the increase is not significant. It could have a potentially positive impact on injury prevention in view of the prospective study of [23]. The latter study focuses on first-year university students studying to become dancers or dance teachers to observe the prevalence of lower limb injuries. The results showed that lower-extremity injuries had the highest one-year incidence (51.4%) and that the main risk factor was a limited ankle dorsiflexion.

In summary, in this study of young recreational dancers, we demonstrated the existence of correlations between kinematic parameters of the grand battement and aesthetic judgment of the execution of this figure by a jury. Moreover, we have shown the short-term effects of dynamic stretching in improving the physical skills related to the execution of the grand battement. It is therefore up to young dancers and dance teachers to incorporate effective muscle stretching methods and to work on improving these kinematic parameters in their training with the subsequent aim of positively influence aesthetic self-expression and judgment of the audience.

## Data Availability

Data are available upon request to the authors.

## Acknowledgments

L.F. thank the students of Mouvements (Tamines, Belgium) and Emergence dance schools (Phillipeville, Belgium) for their spontaneous participation.

F.D., N.R. and F.B. acknowledge financial support of the European Regional Development Fund (Interreg FWVl NOMADe 4.7.360).

